# Efficacy of Chloroquine or Hydroxychloroquine in COVID-19 Patients: A Systematic Review and Meta-Analysis

**DOI:** 10.1101/2020.07.12.20150110

**Authors:** Zakariya Kashour, Muhammad Riaz, Musa A. Garbati, Oweida AlDosary, Haytham Tlayjeh, Dana Gerberi, M. Hassan Murad, M. Rizwan Sohail, Tarek Kashour, Imad M. Tleyjeh

## Abstract

**Background:** Chloroquine (CQ) and hydroxychloroquine (HCQ) show anti-SARS-CoV-2 activity in vitro; however, clinical studies have reported conflicting results. We sought to systematically evaluate the effect of CQ and HCQ with or without azithromycin (AZ) on outcomes of COVID-19 patients.

**Methods:** We searched Medline, Embase, EBM Reviews, Scopus, Web of Science, preprints and grey literature up to July 7, 2020. We included studies that assessed COVID-19 patients treated with CQ or HCQ, with or without AZ. We pooled only adjusted effect estimates of mortality using a random effect model. We summarized the effect of CQ or HCQ on viral clearance and ICU admission/ mechanical ventilation.

**Results:** Out of 1463 citations screened for eligibility, five RCTs and 14 cohort studies were included (20,263 hospitalized patients). Thirteen studies (1 RCT and 12 cohorts) with 15,938 patients examined the effect of HCQ on short term mortality. The pooled adjusted OR was 1.05 (95% CI 0.96-1.15, I^2^=0 %, p=0.647). Six cohort studies examined the effect of HCQ and AZ combination among 14,016 patients. The pooled adjusted OR was 0.93 (95% CI 0.79-1.11, I^2^=59.3%, p=0.003). Two cohort studies and three RCTs found no significant effect of HCQ on viral clearance. One RCT with 48 patients demonstrated improved viral clearance in patients treated with CQ and HCQ. Three cohort studies found that HCQ with or without AZ had no significant effect on mechanical ventilation/ ICU admission.

**Conclusion:** Moderate certainty evidence suggests that HCQ, with or without AZ, lacks efficacy in reducing short-term mortality in patients hospitalized with COVID-19.

**Summary:** This systematic review and meta-analysis showed that in-hospital treatment of COVID-19 patients with antimalarials medications failed to reduce short-term mortality and morbidity with potential harm if used in combination with azithromycin.

## Introduction

The COVID-19 pandemic has claimed hundreds of thousands of human lives and caused enormous economic damage. While the race to develop an effective vaccine continues, repurposing of approved drugs remains the most logical treatment approach for SARS-CoV-2 infection and its complications.

Since the discovery of antiviral effects of Chloroquine (CQ) and hydroxychloroquine (HCQ) more than 50 years ago, interest in exploring their therapeutic potential against various viral infections continued relentlessly [1]. CQ/HCQ have been tested against numerous viruses such as Human deficiency virus-1 (HIV-1), SARS, MERS-CoV, influenza, dengue, Ebola, Zika, chikungunya and other viruses [2-10].

Several mechanisms have been proposed for the anti-SARS-CoV-2 effects of CQ/HCQ. All of which are secondary to their ability to raise intracellular pH which affects particularly the endosome function [11, 12]. CQ/HCQ can interfere with all stages of the viral life cycle [11]. They have the potential to hinder SARS-CoV-2 binding to its cell membrane receptor, ACE2 through their interference with glycosylation process of the ACE2 protein that results in reducing its binding affinity to SARS-CoV-2 virus. CQ/HCQ could also prevent fusion of the viral particles to the host cell membrane and prevent their cell entry. Furthermore, CQ/HCQ can also inhibit viral replication, assembly and release of viral particles from the host cells [11]. CQ/HCQ also alter endosomal antigen processing and modulate both, the innate and adaptive immune responses [11, 12]. This leads to decreased production of pro-inflammatory cytokines like tumor necrosis factor-alpha (TNF-α), and interleukins-1 beta and 6 (IL-1β and IL-6).

Additionally, CQ/HCQ improve endothelial function and reduce prothrombotic state [13]. These properties could have favorable effects in patients with severe COVID-19 disease.

With the emergence of SARS-CoV-2 virus and its rapid spread across the globe, it was natural to test the antiviral effects of CQ/HCQ against this new threatening infection. The enthusiasm for their widespread clinical use in the treatment of COVID-19 disease escalated with the early studies reporting their effective in vitro antiviral effects against SARS-CoV-2 virus [14-16].

An early interim analysis of 100 COVID-19 patients was reported by a group from China where they found that CQ therapy was associated with less severe pneumonia, shorter disease course and faster viral clearance [17]. Another small nonrandomized study of 20 patients from France revealed reduced nasopharyngeal viral carrier sate at 6 days after the initiation of treatment with HCQ and azithromycin (AZ) [18]. These limited data along with the political support for CQ/HCQ use led clinicians worldwide to use them indiscriminately and to include them in their institutional protocols and guidelines for the treatment of COVID-19 disease as a monotherapy or in combination with AZ. This rapid adoption of CQ/HCQ was associated with astronomical increase in CQ/HCQ prescription of approximately 2000% [19].

While numerous large randomized clinical trials (RCTs) were started in different countries worldwide, several observational studies addressing the efficacy and safety of CQ/HCQ in the treatment of COVID-19 disease got published along with preliminary results from some RCTs. These studies have different methodologies and sample sizes, and produced mixed results ranging from reduced mortality and improved other clinical outcomes to increased mortality among COVID-19 patients.

The absence of robust clinical evidence for their efficacy and the potential serious drug induced adverse events associated with CQ/HCQ use, calls for rigorously conducted systematic reviews/meta-analyses of the available clinical data to present a clearer picture about their efficacy and provide data-informed view regarding their utility in the treatment of COVID-19. In this study, we set to perform a systematically review and meta-analysis of the literature regarding the efficacy of CQ or HCQ in patients with COVID-19.

## Methods

### Inclusion and Exclusion Criteria

We followed Preferred Reporting Items for Systematic Reviews and Meta-analyses (PRISMA)[20] guidelines for reporting systematic review and meta-analysis of observational studies guidelines. We included 1) RCTs or 2) cohorts or case control studies reporting on adjusted effect estimates of the association between Hydroxychloroquine and the following endpoints: (1) short term mortality, (2) mechanical ventilation/ ICU admission and (3) viral clearance.

### Literature Search

The literature was searched by a medical librarian for the concepts of CQ or HCQ combined with COVID-19. The search strategies were created using a combination of keywords and standardized index terms. Searches were run through July 7, 2020 in Ovid EBM Reviews, Ovid Embase (1974+), Ovid Medline (1946+ including epub ahead of print, in-process & other non-indexed citations), Scopus (1970+) and Web of Science (1975+). Search strategies are provided in the supplement table 1. We also searched for unpublished manuscripts using the medRxiv services operated by Cold Spring Harbor Laboratory and Research Square preprints. In addition, we searched Google Scholar and the references of eligible studies and review articles.

Two reviewers independently identified eligible studies (ZK and OA) and 4 reviewers (ZK, MG, OA, HT) extracted the data into a pre-specified data collection form. A senior reviewer verified all data included in the analyses (IT).

Two reviewers (ZK and OA) independently assessed risk of bias for each study using the (RoB 2) of the Cochrane risk-of-bias tool for randomized trials [21] and the Newcastle–Ottawa instrument for cohort studies and case–control studies [22]. Reviewers judged each criterion for risk of bias and resolved any disagreements by discussion with a third senior reviewer. We assessed the certainty of evidence for each of our outcomes using the GRADE (Grading of Recommendations Assessment, Development, and Evaluation) approach [23, 24]. This method evaluates the certainty of evidence by assessing the following domains: Limitations, indirectness, inconsistency, imprecision, and publication bias.

### Statistical analysis

We pooled studies using the DerSimonian-Laird random-effects model (and constructed corresponding forest plots). Pooled adjusted effect estimates (ORs and HRs) were obtained by combining the estimates of log (adjusted effect estimate) from each study. Endpoints that we considered a priori for the meta-analysis were: (1) short term mortality, (2) mechanical ventilation/ ICU admission and (3) viral clearance. We evaluated heterogeneity using the I^2^ statistic which estimates the variability percentage in effect estimates that is due to heterogeneity rather than to chance—the larger the I^2^, the greater the heterogeneity. We conducted sensitivity analyses to assess the impact of (1) risk of bias in included studies, and (2) the selection of study population (general populations vs. specific populations), on the overall estimate of effect. We constructed funnel plots and performed an Egger precision-weighted linear regression test as a statistical test of funnel plot asymmetry and publication bias. All analyses were conducted using Stata version 16 statistical software (StataCorp, College Station, Texas).

## Results

### Included studies

Out of 1,896 papers screened for eligibility, 5 RCTs [25-29], 14 cohort studies [30-43] were included (Figure 1) with a total of 20,263 patients. Characteristics of included studies are described in Tables 1 and 2. Characteristics of the patients in each study can be found in Supplementary Table 5. All studies reported on patients hospitalized with COVID-19. Alberici et al. also included out-patients (39%). Most of the studies included patients presenting with varying levels of severity; Ip et al. [33], Magagnoli et al. [35], Rosenberg et al. [39] included patients who were hypoxic, t or were admitted to the ICU, in addition to patients with relatively more stable vitals. Paccoud et al. [38] included patients with varying pneumonia severity. Yu et al. only studied patients with critical disease whereas Geleris et al. [32] and Mahevas et al. [36] excluded them.

**Table 1.**
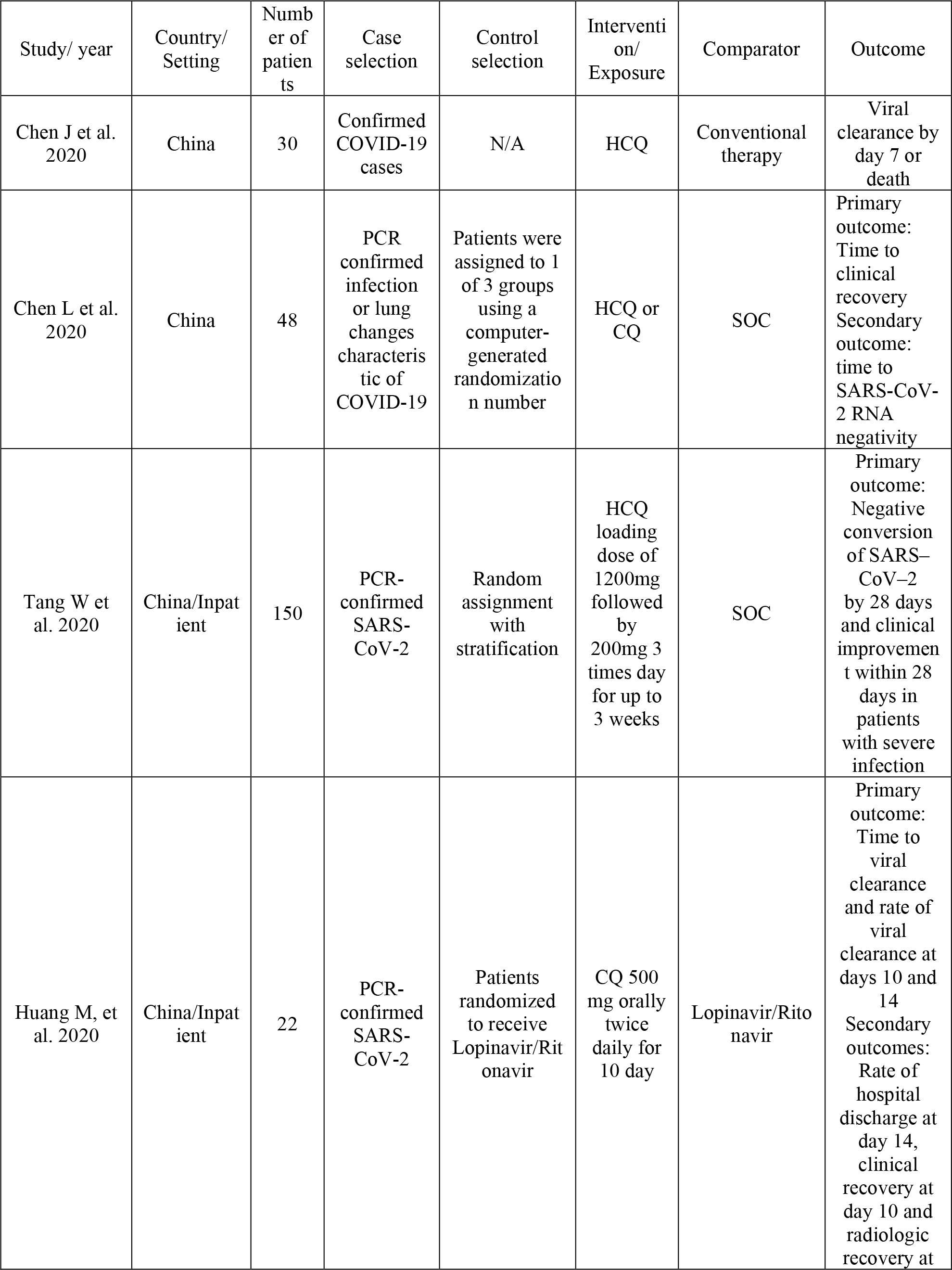
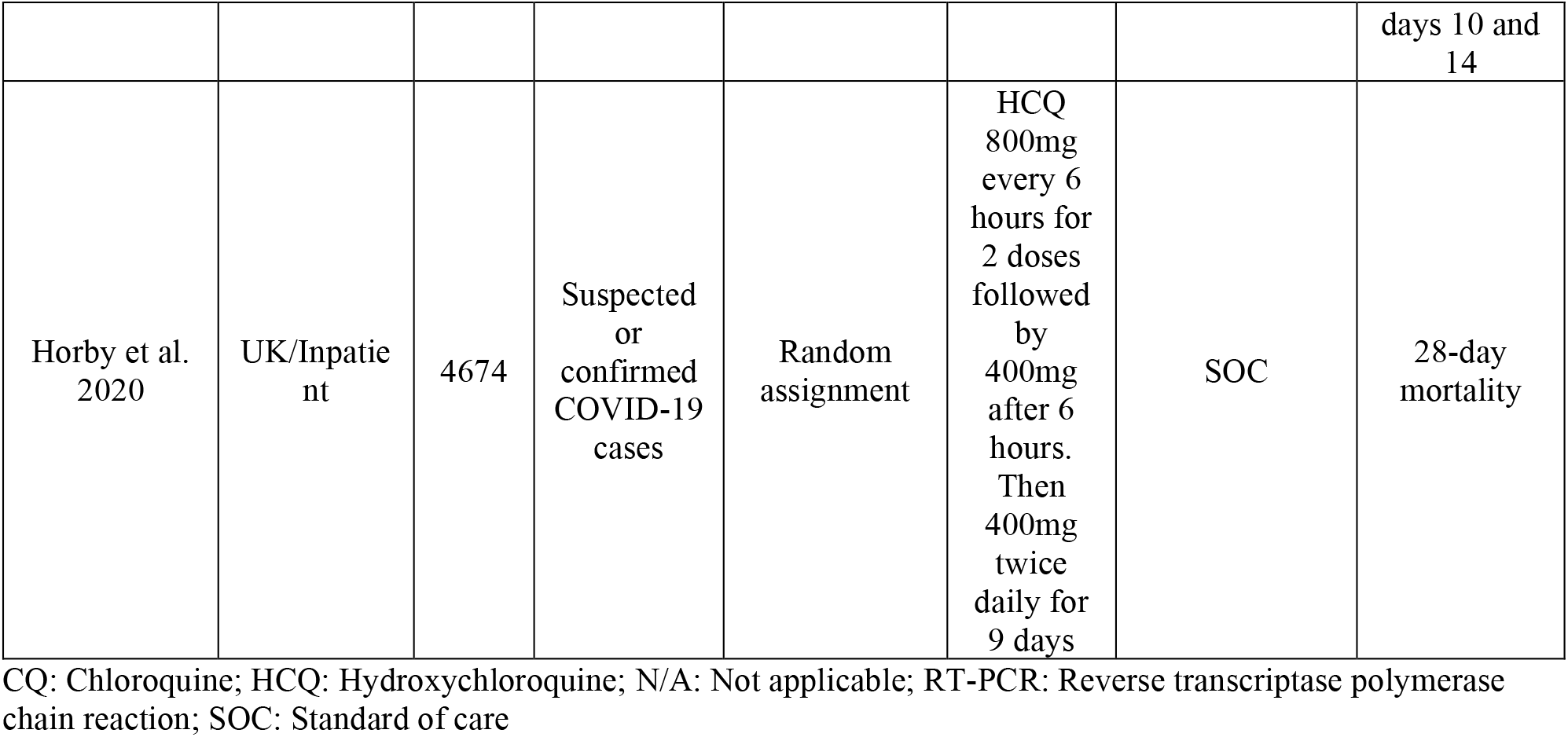
Characteristics of Randomized Clinical Trials.

| Study/ year | Country/<br>Setting | Number of<br>patients | Case<br>selection | Control<br>selection | Intervention/<br>Exposure | Comparator | Outcome |
| --- | --- | --- | --- | --- | --- | --- | --- |
| Chen J et al.<br>2020 | China | 30 | Confirmed<br>COVID-19<br>cases | N/A | HCQ | Conventional<br>therapy | Viral<br>clearance by<br>day 7 or<br>death |
| Chen L et al.<br>2020 | China | 48 | PCR<br>confirmed<br>infection<br>or lung<br>changes<br>characteristic<br>of<br>COVID-19 | Patients were<br>assigned to 1<br>of 3 groups<br>using a<br>computer-<br>generated<br>randomization<br>number | HCQ or<br>CQ | SOC | Primary<br>outcome:<br>Time to<br>clinical<br>recovery<br>Secondary<br>outcome:<br>time to<br>SARS-CoV-2 RNA<br>negativity |
| Tang W et<br>al. 2020 | China/Inpatient | 150 | PCR-confirmed<br>SARS-CoV-2 | Random<br>assignment<br>with<br>stratification | HCQ<br>loading<br>dose of<br>1200mg<br>followed<br>by<br>200mg 3<br>times day<br>for up to<br>3 weeks | SOC | Primary<br>outcome:<br>Negative<br>conversion<br>of SARS-CoV-2<br>by 28 days<br>and clinical<br>improvement<br>within 28<br>days in<br>patients<br>with severe<br>infection |
| Huang M, et<br>al. 2020 | China/Inpatient | 22 | PCR-confirmed<br>SARS-CoV-2 | Patients<br>randomized<br>to receive<br>Lopinavir/Ritonavir | CQ 500<br>mg orally<br>twice<br>daily for<br>10 day | Lopinavir/Ritonavir | Primary<br>outcome:<br>Time to<br>viral<br>clearance<br>and rate of<br>viral<br>clearance at<br>days 10 and<br>14<br>Secondary<br>outcomes:<br>Rate of<br>hospital<br>discharge at<br>day 14,<br>clinical<br>recovery at<br>day 10 and<br>radiologic<br>recovery at days 10 and 14 |
| Horby et al. 2020 | UK/Inpatient | 4674 | Suspected or confirmed COVID-19 cases | Random assignment | HCQ 800mg every 6 hours for 2 doses followed by 400mg after 6 hours. Then 400mg twice daily for 9 days | SOC | 28-day mortality |

CQ: Chloroquine; HCQ: Hydroxychloroquine; N/A: Not applicable; RT-PCR: Reverse transcriptase polymerase chain reaction; SOC: Standard of care

**Table 2.**
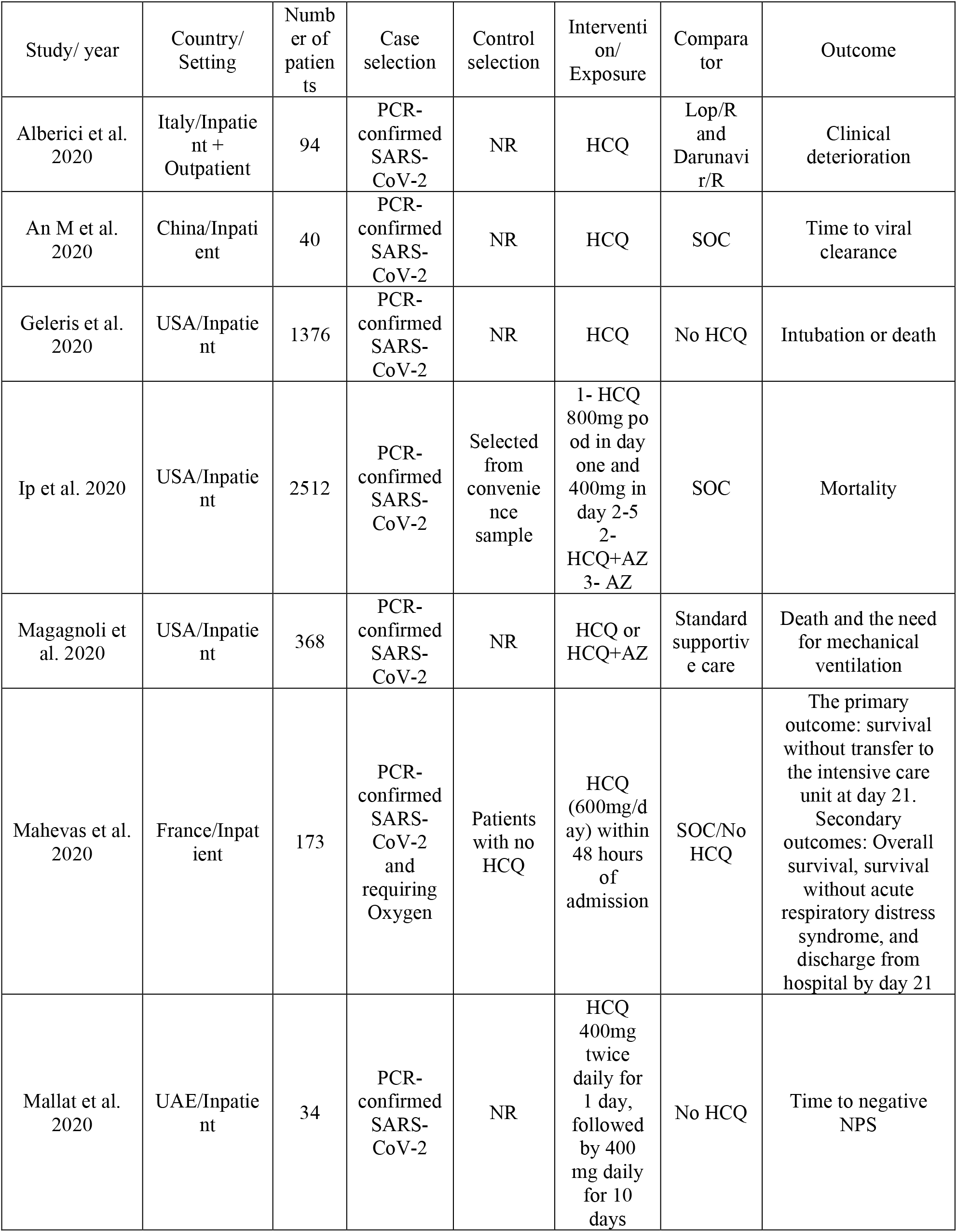
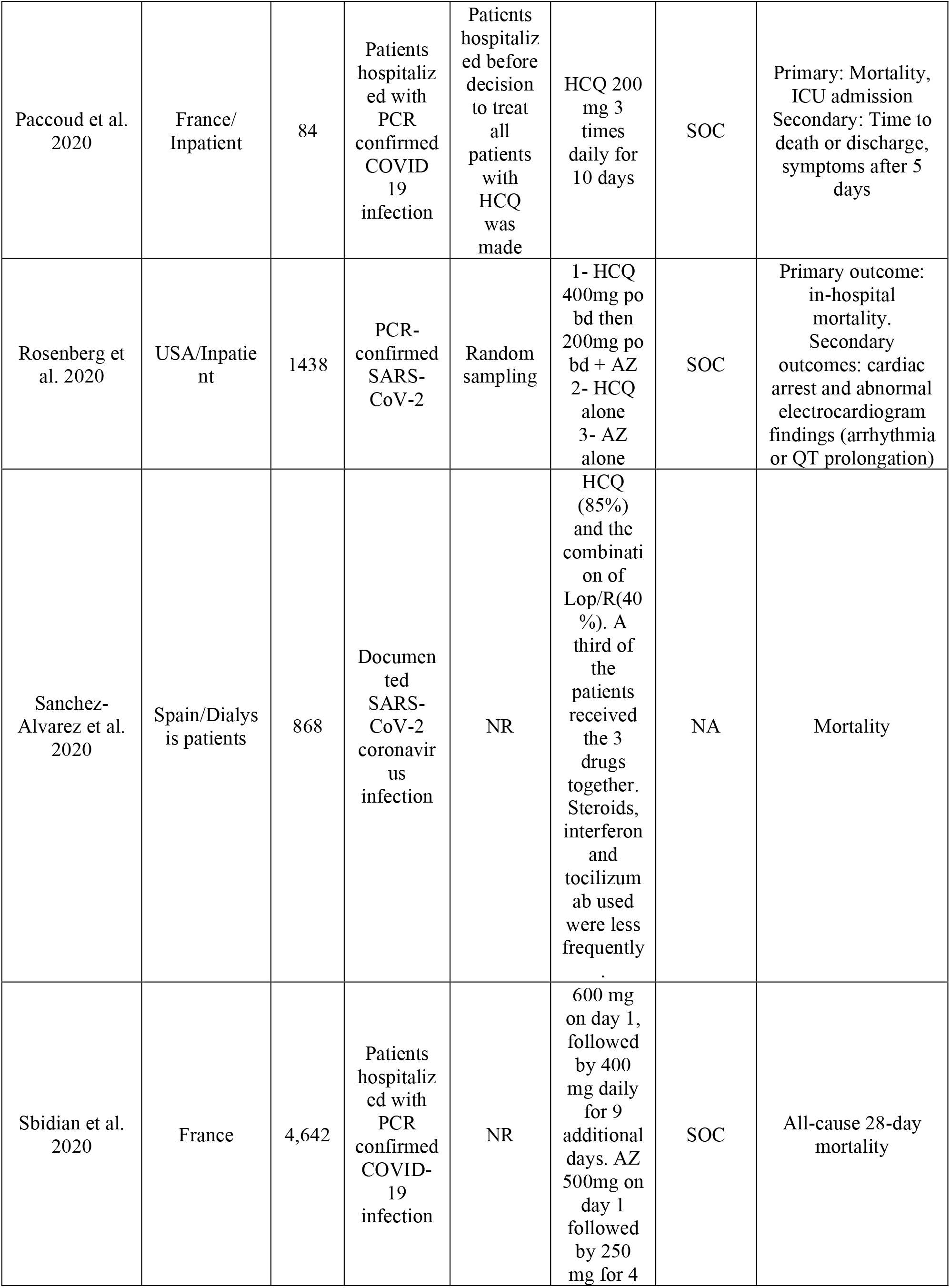
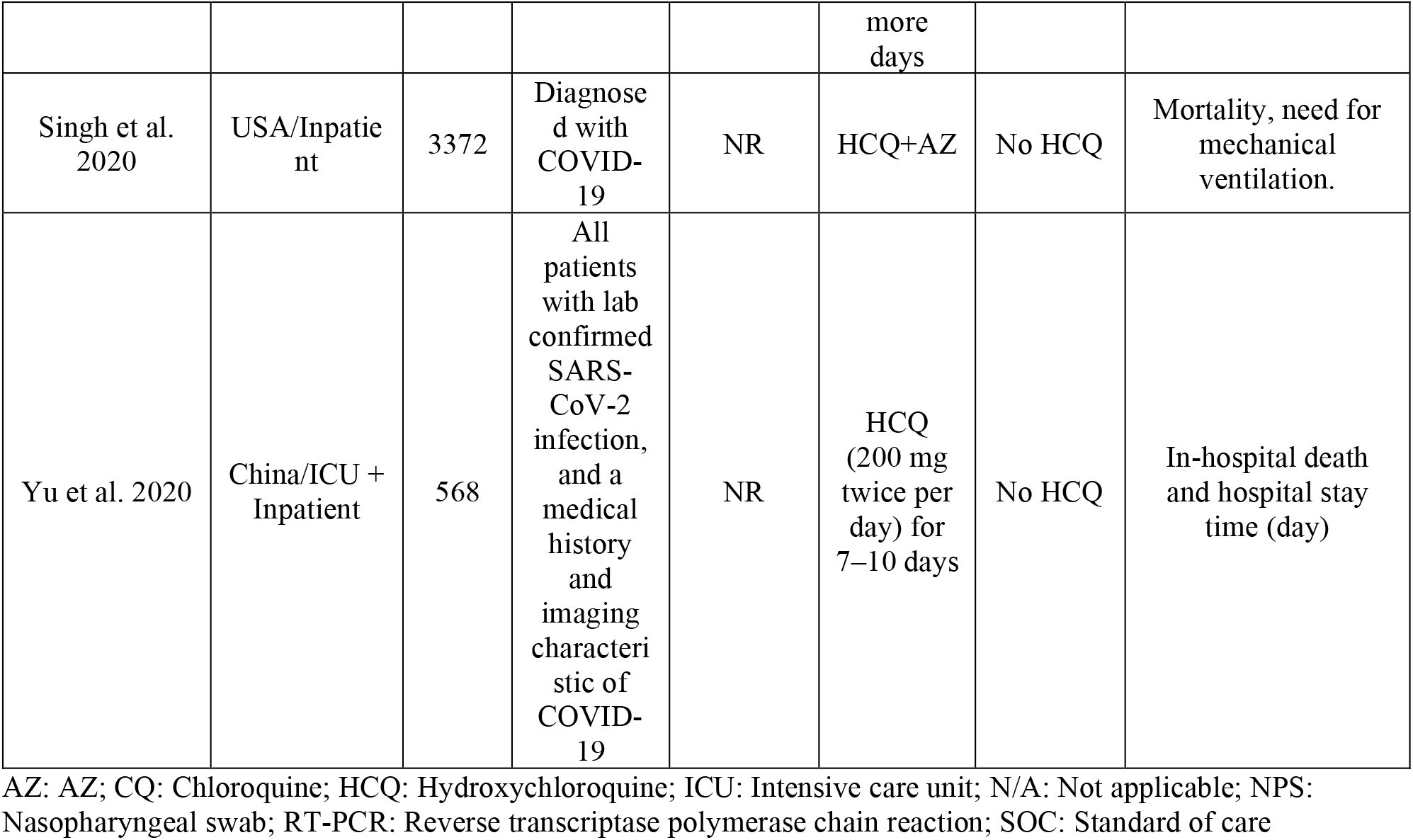
Characteristics of Cohort studies.

**Figure 1.**
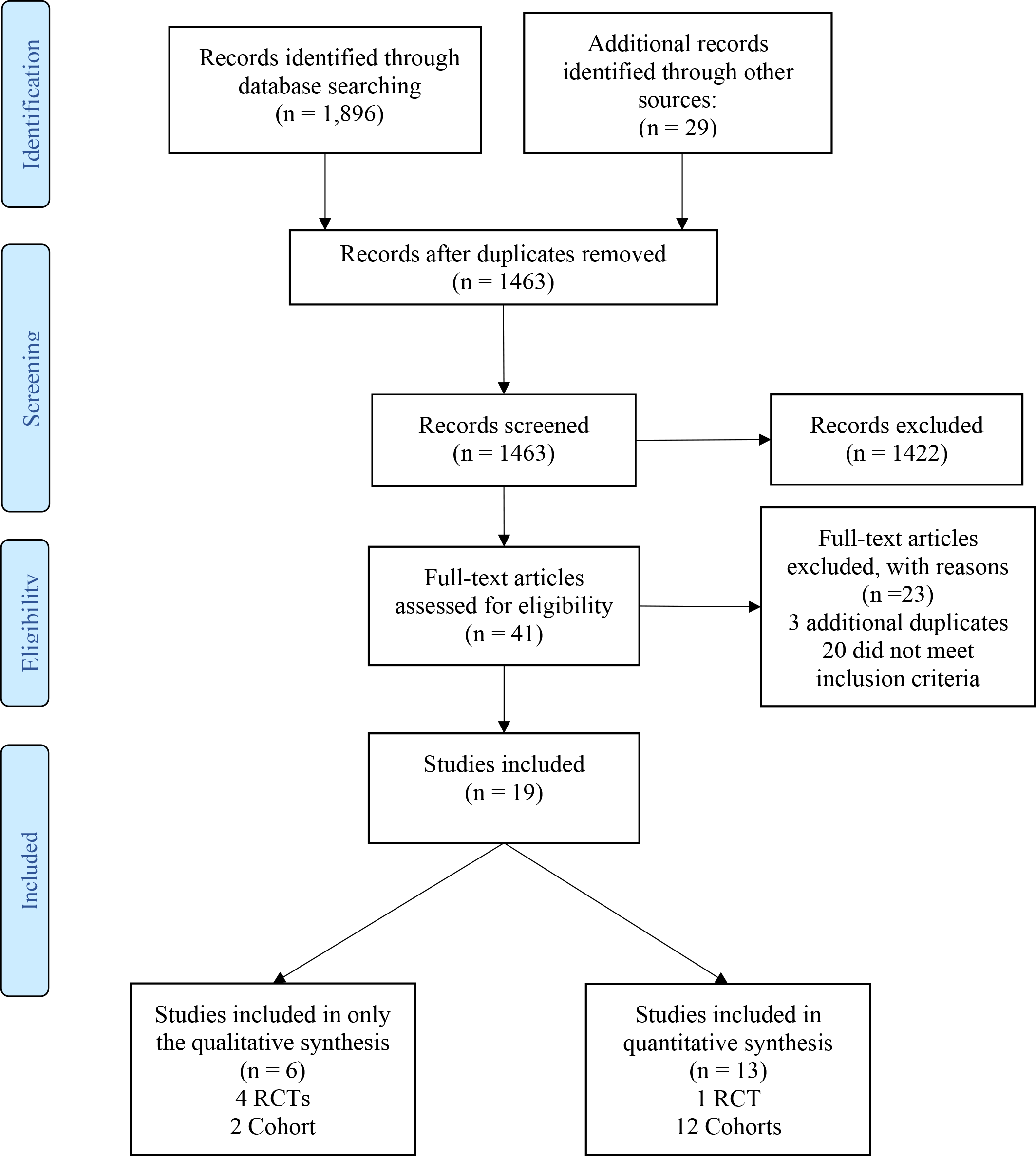
PRISMA Flow Diagram of Eligible Studies

**Figure 2.**
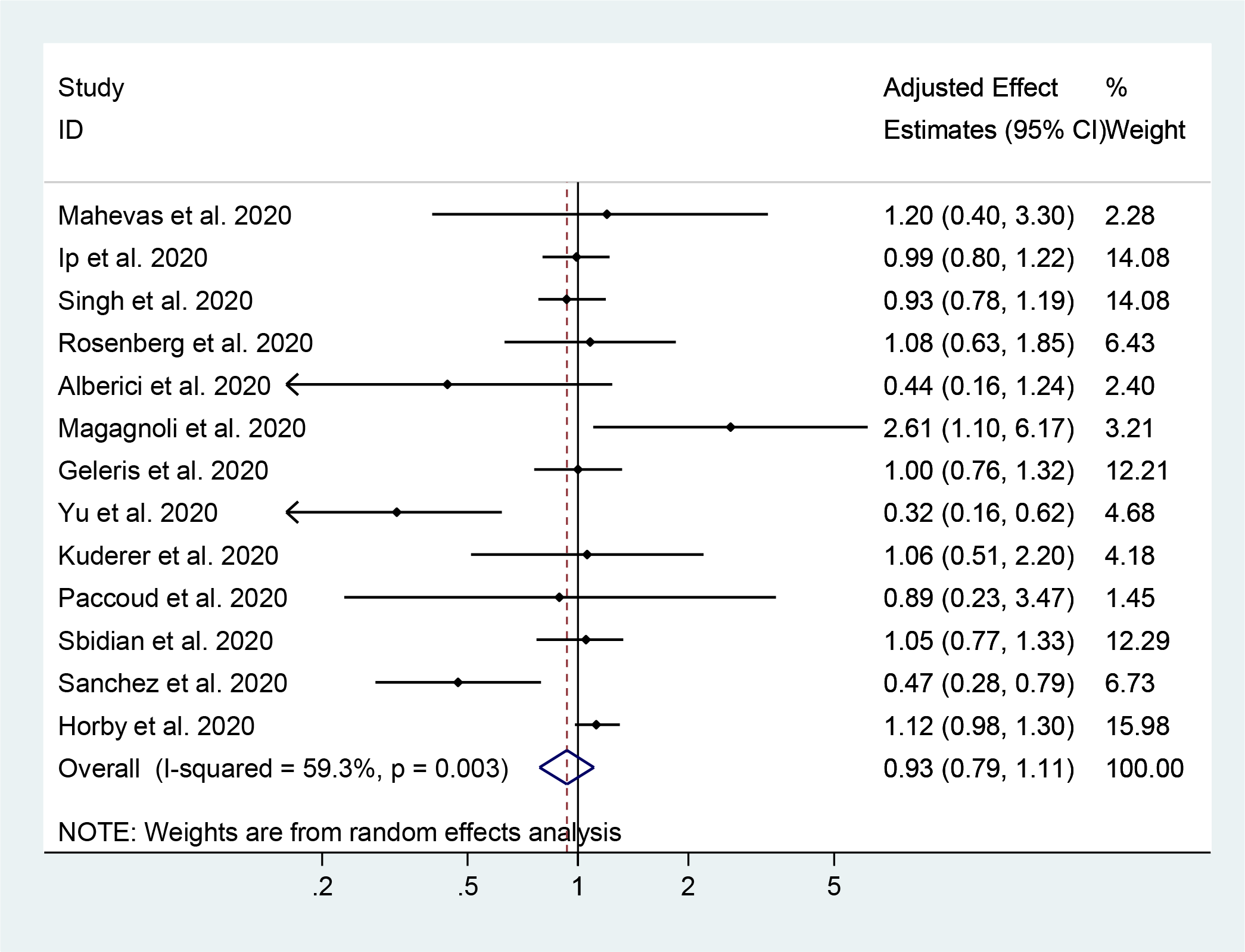
Association between Hydroxychloroquine and short-term mortality in COVID-19 patients: (All cohorts and 1 RCT)

**Figure 3.**
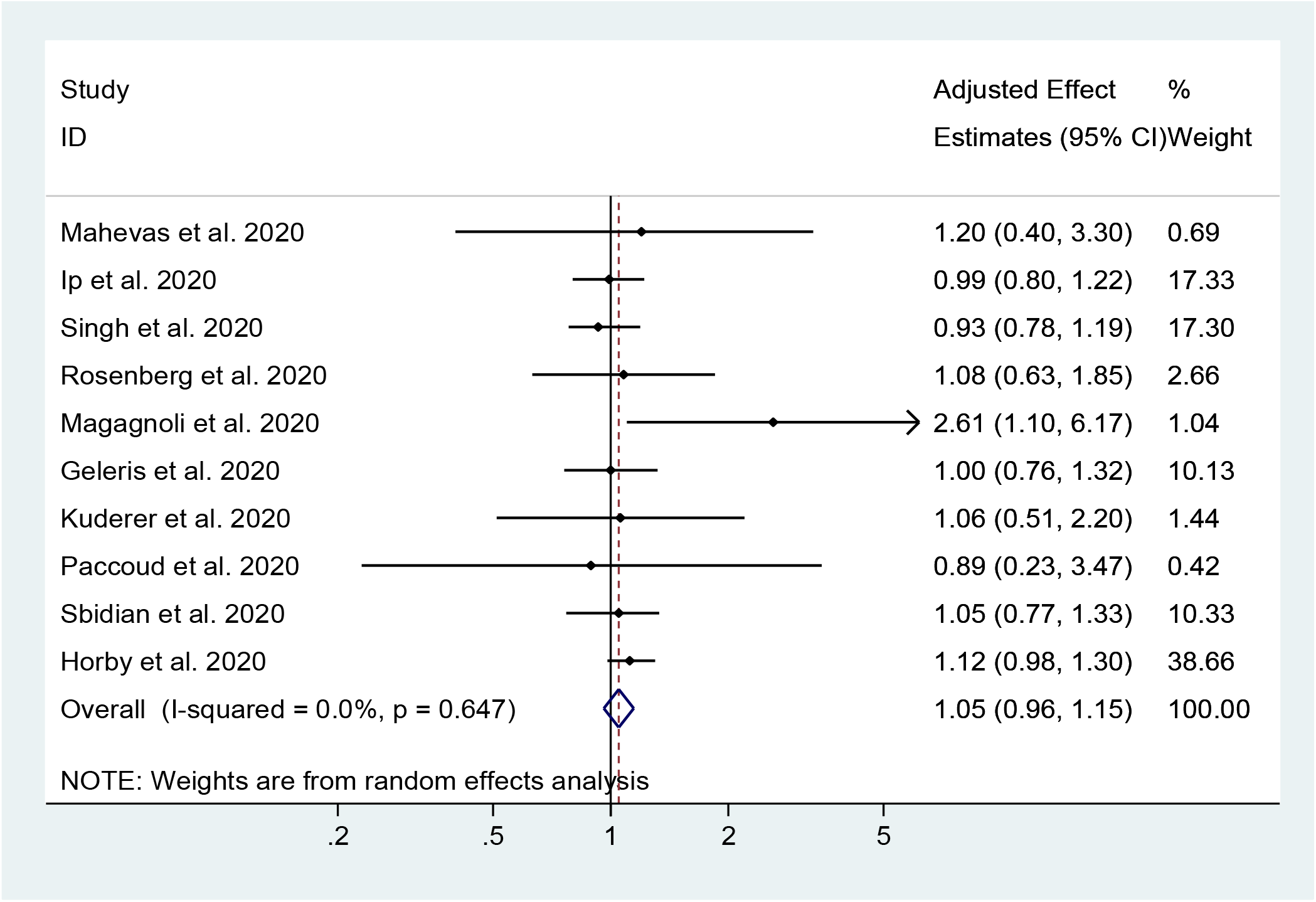
Association between Hydroxychloroquine and short-term mortality in COVID-19 patients: (Excluding studies at high risk for bias)

**Figure 4.**
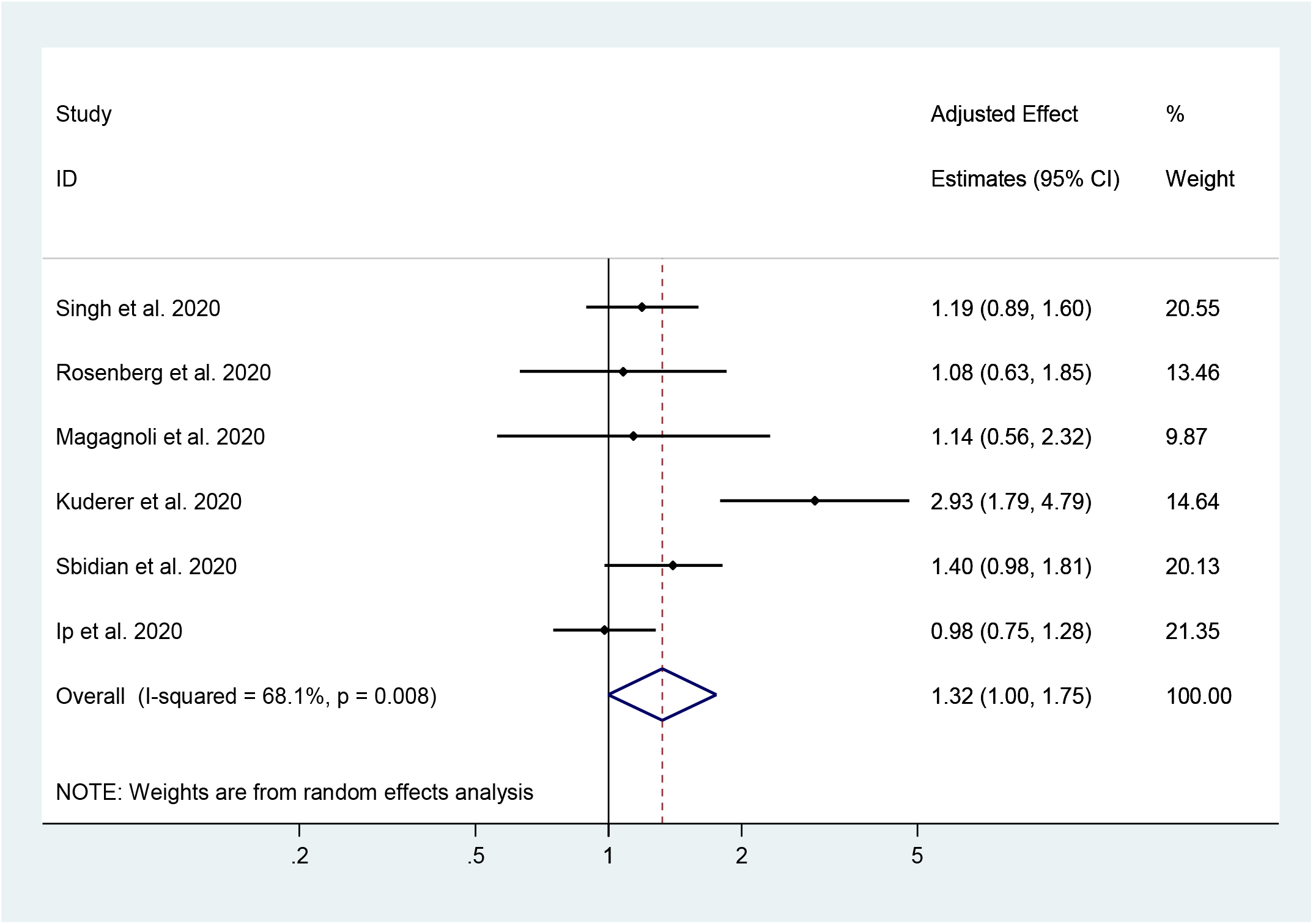
Association between Hydroxychloroquine and AZ combination and short-term mortality in COVID-19 patients

The quality of the observational studies was assessed using the Newcastle-Ottawa Scale (Supplementary Table 11). With regards to patient selection, all but four studies had adequate representation of a general population with COVID-19 infection. Alvarez et al. [31] and Alberici et al.[30] studied hemodialysis patients, Yu et al. [42] studied patients with severe COVID-19 infection, and Kuderer et al. [34] studied patients with cancer. With regards to comparability, three studies [30, 31, 37] did not adequately adjust for confounders in their analyses. Given the relatively short course of disease, all studies were considered to have had a satisfactory follow-up duration. Three studies [30, 31, 42] that assessed mortality were considered at high risk of bias, and a sensitivity analysis was performed by excluding these studies to assess heterogeneity. The results of the Cochrane ROB assessment for RCTs are found in supplementary table 12. Of the 5 included RCTs 3 were considered high risk [27-29], one RCT was low risk [25], and one had no available manuscript for quality assessment [26].

Some studies were initially included in our review as eligible studies but were later excluded for various reasons. Chen Z et al. [44] reported time to clinical recovery and changes in radiologic parameters. Feng et al. [45] reported disease progression in patients receiving chloroquine, whereas only Esper [46] reported hospitalization in outpatients taking HCQ compared to controls (1.9 vs 5.4 P<0.0001). Four studies were excluded over serious methodology concerns [47-50] and one study was retracted due to concerns over the validity of the patient information [51]. GRADE was used to assess the certainty of evidence for each outcome. The findings were then summarized, along with the overall certainty of evidence (Table 3).

**Table 3.** Summary of Outcomes, key findings, and certainty of evidence.

| Treatment | Outcome | Study design:<br>No. of studies | Findings and Magnitude of Effect | Strength of evidence |
| --- | --- | --- | --- | --- |
| HCQ | Mortality | RCT:1<br>Cohort:12 | 1. Studies with moderate and high risk of bias, with consistent but imprecise EE's found no significant association between HCQ and mortality EE's ranged from 0.32 (0.16- 0.62) to 2.61 (1.10-6.17)<br>2. Pooled adjusted OR from 9 cohort studies at moderate risk of bias and 1 RCT at low risk of bias found no significant association between HCQ and mortality: 1.05 (95% CI 0.96-1.15 $I^2=0\%$, $p=0.647$), with no heterogeneity or evidence of publication bias | Moderate (No effect) |
|  | Viral Clearance | RCTs: 3<br>Cohort: 2 | Studies with Low and high risk of bias, inconsistent and imprecise EE's found no association between HCQ and viral clearance. EE's ranged from: 0.46 (95% CI 0.04- 5.75) to 5.68 (95% CI 1.05-10.08). | Very low |
|  | Mechanical ventilation/ ICU admission | Cohort: 3 | Studies with moderate risk of bias, consistent and precise results found no significant association between HCQ and the composite outcome EE's ranged from 0.81 (95% CI 0.55- 1.18) to 1.43 (95% CI 0.53-3.79) | Very low |
| HCQ+AZ | Mortality | Cohort:6 | 1. Studies with moderate risk of bias showed a trend towards increased mortality. AEE ranged from 0.98 (0.75- 1.28) to 2.93 (1.79- 4.79)<br>2. Pooled adjusted OR=1.15, 95% CI 0.99-1.34, $I^2=0.0\%$) from 5 cohort studies at moderate risk for bias. | Low (Higher mortality) |
|  | Mechanical ventilation/ ICU admission | Cohort: 2 | Studies with moderate risk of bias, consistent and precise results found no significant association between HCQ+AZ and the composite outcome | Very low |
| CQ | Viral Clearance | RCTs: 2 | 1 RCT with high risk of bias, in consistent and imprecise EE's showed no significant effect of Chloroquine on viral clearance EE:1.07 (0.44,2.56). 1 RCT with high risk of bias demonstrated shorted time to viral clearance in the CQ group (Median 2.5 days IQR 2-3.8) vs control group (Median 7 days IQR 2-10) | Very low |

AEE: Adjusted effect estimate; AZ: AZ; CI: Confidence interval CQ: Chloroquine EE: Effect estimate; HCQ: Hydroxychloroquine; RCT: Randomized clinical trial

### Treatment efficacy

#### Mortality

A total of 13 studies[26, 30, 31, 33-36, 38-42] (1 RCT and 12 cohorts) with 19,573 patients examined the effect of HCQ on short term mortality in hospitalized COVID-19 patients. The pooled adjusted OR was 0.93 (95% CI 0.79-1.11, I^2^=59.3%, p=0.003) indicating no significant association between HCQ and mortality. There was moderate heterogeneity among the included studies. In a sensitivity analysis, after excluding the 3 studies with high risk of bias [30, 31, 42] the pooled adjusted OR was 1.05 (95% CI 0.96-1.15, I^2^=0 %, p=0.647).

A total of 6 cohort studies [33-35, 39-41] with 3430 patients examined the effect of HCQ and AZ combination on mortality. The pooled adjusted OR was 1.32 (95% 1.00-1.75, I^2^=68.1% p=0.008) with a higher odds for mortality in the combination therapy group compared with the control group. Excluding the study of Kuderer et al. [34], that included only patients with cancers, eliminated this heterogeneity (adjusted OR=1.15, 95% CI 0.99-1.34, I^2^=0.0%). There was no publication bias on visual inspection of funnel plots (Supplementary Figures 2-5). Additionally, Egger’s regression did not detect any significant publication bias (p= 0.276).

#### Viral Clearance

Three RCTs (2 with high risk of bias [28, 29] and one with low risk [25] of bias assessed using Cochrane ROB scale for quality assessment of RCTs) and two cohort study [37, 43] (with a high risk of bias assessed using NOS) assessed the effect of HCQ on viral clearance. One RCT [29] demonstrated significant improvement in time to viral clearance (2.0 days IQR: 2.0-3.5); while the other two didn’t show any significant effect on time to viral clearance: 0.46 (95% CI 0.04-5.75) and 0.846 (95% CI 0.58-1.23) respectively. Neither cohort studies demonstrated an association between HCQ and slower viral clearance adjusted OR 5.68 (95% CI 1.05-10.08) [37] and adjusted HR1.53 (95% CI 0.83-2.94)[43] respectively.

Two RCTs [27, 29] with high risk of bias studied the effect of chloroquine on viral clearance. In one study with 22 patients [27], there was no significant effect on viral clearance OR 1.07 (95% CI 0.44-2.56). Another RCT with 48 patients found a significant difference in time to viral clearance in the CQ group compared with the control group 2.5 days (IQR: 2.0-3.8) and, vs 7.0 days (IQR: 3.0-10.0) respectively[29].

Six studies in our meta-analysis evaluated the effect of therapy on viral clearance; of which five had a high risk of bias and thus effect estimates were not pooled together.

#### Mechanical ventilation/ ICU admission

Three cohort studies assessed the association between HCQ and the composite outcome of mechanical ventilation or ICU admission [35, 36, 41]. None of the studies found any association between HCQ and the composite outcome [1.1 (95% CI 0.476-2.5), 1.43 (95% CI 0.53-3.79) and 0.81 (95% CI 0.55-1.18) respectively].

Additionally, two cohorts [35, 41] failed to demonstrate any significant association between HCQ+AZ and the composite outcome of mechanical ventilation or ICU admission AEE 0.43 (95% CI 0.16-1.12) and 0.976 (95% CI 0.64-1.49) respectively.

## Discussion

### Main findings

Our systematic review and meta-analysis included 12 cohorts and 1 RCT, which addressed the association between HCQ therapy in improving and mortality in patients hospitalized with COVID-19 disease. Among a total of 19573 patients, we found with moderate level of certainty, that HCQ monotherapy did not reduce short-term mortality among COVID-19 patients, which remained statistically non-significant even after excluding three studies at high risk of bias. These observations did not change when we included in the model only the cohort studies. Moreover, we found that the use of combination of HCQ and AZ was associated with a trend of increased mortality. The study by Kruderer et al. analyzed cancer patients and demonstrated higher mortality than other studies creating significant heterogeneity. Excluding this study did not decrease the risk of mortality. Because of the limited number of studies and/or high risk of bias, we could not meta-analyze other clinical outcomes such as viral clearance, risk of ICU admission, and need for mechanical ventilation.

Our findings of lack of efficacy of HCQ in the inpatient clinical setting despite its effective in vitro inhibitory actions against SARS-CoV-2 virus is consistent with previous observations with other viral illnesses. Numerous studies demonstrated significant in vitro inhibitory effects of CQ/HCQ against coronaviruses and non-coronaviruses [11]. For example, CQ at half-maximum effective concentration (EC_50_) of 8.8 ± 1.2 µM effectively inhibited SARS-CoV replication in Vero E6 cells [3]. CQ was also shown to inhibit MERS-CoV and alphacoronavirus HCoV-229E replication in vitro in a dose dependent manner [4]. Likewise, CQ/HCQ exhibited in vitro inhibitory effects against several other viruses such as HIV-1, influenza, dengue, Ebola, Zika, chikungunya viruses and others [2, 5-10]. Although CQ/HCQ have shown consistent broad spectrum in vitro antiviral effects, their in-vivo and clinical antiviral effects were disappointing. For instance, CQ was ineffective in preventing or ameliorating influenza following viral challenge in mouse and ferret models [5] and did not prevent influenza infection in a randomized, double-blind placebo-controlled trial in humans [52]. CQ also resulted in worse outcomes in a guinea pig model of Ebola infection [7] and was shown to enhance chikungunya viral infections in different animal models including non-human primates [53]. Moreover, CQ was s ineffective in improving the course of chikungunya in humans [53, 54]. CQ was also tested in a randomized controlled trial of 307 patients with dengue virus and failed to reduce duration of viremia or NS1 antigenemia [55]. In the case of HIV-1, the use of CQ/HCQ was inconclusive and hence they were not endorsed for routine use in the treatment of HIV-1 infection [56].

### Possible reasons for lack of efficacy of HCQ in the treatment of COVID-19 disease

The discrepancy between the observed in vitro anti-SARS-CoV-2 effects of CQ/HCQ and the lack of efficacy in clinical studies, which mirrors previous observations with other viral infections could be due to three main reasons:

First, most of in vitro studies employ pre-treatment protocols where cells are treated with the drugs before infecting them with the tested virus. In vitro studies that compared pre-treatment and post-infection treatment have shown that CQ/HCQ have less effective antiviral activities if added after infection [4, 16, 57]. For example, Vincent et al. [57] showed that e CQ at 0.1, 1.0 and 10 µM added 20-24 hours before the infection with SARS-CoV decreased infectivity by 28%, 53% and 100%. However, if CQ is added 3-5 hours after infecting the cells, higher concentrations of CQ of up to 50 µM were needed to decrease infectivity [57]. This may raise the possibility that chronic or prophylactic use of CQ/HCQ may reduce the risk of acquiring SARS-CoV-2 infection. However, a recent large population study of 14,250 individuals showed that chronic use of HCQ was not protective against SARS-CoV-2 infection [58].

Second, a wide range of EC_50_ values were reported for CQ and HCQ and in the case of SARS-CoV-2, the EC_50_ for CQ ranges between 1.13 and 7.36 µM and between 0.72 and 17.31 µM for HCQ [11]. It is worth noting that the lowest EC_50_ of 0.72 µM for HCQ was reported by Yao et al in their post-infection experiments, was different from the lowest EC_50_ of 5.85 µM in their pre-treatment experiments [16]. None of the other investigators reported such lower EC_50_ for CQ of HCQ with SARS-CoV-2 or any other viruses. Achieving adequate blood and tissue drug concertation is essential for proper antiviral activity In mice, a high dose of 90 mg/kg twice a day of CQ was necessary to achieve a steady state blood levels of 2.5 µg/ml [8]. Considering high doses of CO/HCQ in COVID-19 patients can be associated with increased adverse events as shown in a recent RCT where high dose CQ has been shown to be associated with significant toxicity in COVID-19 patients [59]. In addition, it is not clear what is the optimal CQ or HCQ blood levels for effective antiviral action since the suggested levels were based on widely differing in vitro EC_50_ estimates. In a study of 40 HIV-1 patients treated with HCQ 800 mg/day for 8 weeks, the range of HCQ blood concentration was 0.27-1.0 µg/ml [60]. Only those HIV-1 patients who achieved the highest HCQ blood concentrations had favorable response to HCQ [60]. There are only two small studies that looked at the pharmacokinetics of HCQ in COVID-19 patients [18, 60, 61]. In the first study Gautret et al. determined the mean HCQ levels in 20 patients treated with HCQ 600 mg/day and was 0.46 µg/ml [18]. This blood concentration is lower than the lowest estimated levels of 0.48 µg/ml based on the lowest effective in vitro concentration of 0.72 µM. Perinel et al showed that only 61% of 13 patients treated with HCQ 600 mg/day achieved what they considered the minimum therapeutic concentration of 1 µg/ml with the mean time to reach this concentration of 2.7 days [61]. Based on published data and their own, Balevic et al. found that the average serum/plasma HCQ concentration were below the lowest antiviral target levels for SARS-CoV-2 of 0.48 µg/ml in all studies [62]. These studies indicate that current HCQ dosing is probably suboptimal to achieve adequate blood levels necessary for effective antiviral activity.

Third, Antimalarials exhibit anti-inflammatory and immunomodulatory effects where they can decrease the production of pro-inflammatory cytokines, and improve endothelial function and reduce prothrombotic state [12, 13]. These effects would be very beneficial in patients with severe COVID-19 disease; however, HCQ reduce the affinity of toll-like receptor 7 and 9 (TLR7 and TLR9) to viral RNA and also inhibit cyclic GMP-AMP synthase (cGAS) pathway and hence it inhibits type I interferon response which is the first defense line of the innate immune system against viral infections [12]. This effect might counteract the direct antiviral effects of HCQ and reduce its efficacy in treating COVID-19 disease.

Further research is needed to address these important issues to improve the clinical utility of CQ/HCQ for the treatment of COVID-19 disease, which should include exploring alternate administration routes like intra nasal application and inhalation therapy.

### Safety of CQ/HCQ in the context of COVID-19 disease

Our meta-analysis not only revealed lack of efficacy of HCQ in improving the outcomes of COVID-19 patients but also suggests possible increased risk of mortality when used in combination with AZ. Several studies have shown increased risk of cardiac toxicity among COVID-19 patients treated with CQ/HCQ. Our group have recently conducted a meta-analysis on CQ/HCQ induced cardiac toxicity in COVID-19 patients, which revealed [63] increased risk of QTc prolongation and discontinuation of drug due to QT prolongation. In addition, CQ/HCQ was associated with a clinically significant risk malignant arrhythmias and cardiac arrest [59]. It has also been a common practice to use HCQ in combination with AZ for COVID-19 during the current pandemic. AZ has s been linked to increased risk of sudden cardiac death [63, 64]. Hence, the concomitant use of CQ/HCQ and AZ or other QT prolonging agents could potentially increase the risk of serious cardiac arrhythmias and death. Increased risk of 30-day cardiac death, angina and heart failure complications associated with the combination therapy of HCQ and AZ has also been reported in a recent preprint of a large population study of 323,122 patients [64]. Our findings are consistent with the IDSA recommendations on the use of CQ/HCQ in COVID-19 disease [65], and the recent systematic reviews [62-64]. However, our study has several advantages over the previous reviews (supplementary table 13). First, it is the only study that included both qualitative and quantitative analyses. Second, it has the largest number of identified studies and therefore patient population because our search is the most up to date. Third, in contrast to all previous studies, we included only studies that reported adjusted effect estimates and therefore we avoided including studies at high risk of bias due to confounding. Fourth, we provided adequate assessment of the certainty of evidence using the GRADE classification. Finally, we offered a comprehensive discussion regarding the probable mechanisms of lack of efficacy of HCQ in COVID-19, which will inform and stimulate further research in this area.

## Strengths and Limitations

This meta-analysis has several strengths. Firstly, published and unpublished studies were included, which reduces publication bias. We also employed rigorous methodologies where we excluded studies that did not report adjusted odds or hazard ratios and those with poor methodology. We analyzed and reported monotherapy and combination therapy separately. We also examined mortality and other clinical outcomes separately and performed sensitivity analyses to eliminate sources of between studies heterogeneity. However, our study has several limitations; all of our included studies except one were observational studies which are prone to bias; including confounding by allocation, survival bias and residual confounding. Our group and others have shown that survivor bias, which occurs because patients who live longer are more likely to receive treatment than those who die early, could change associations from benefit to harm [66, 67]. Moreover, as with all observational studies, residual confounding could inflict any observed association [68] even with appropriate adjustment or propensity score matching. Nevertheless, the direction of these biases is supposed to be in favor of hydroxychloroquine efficacy. In addition, our pooled estimates are consistent with the results of the interim report from the RECOVERY trial which lends support to our findings.

## Conclusion

This systematic review and meta-analysis indicate with moderate level of certainty, that hydroxychloroquine monotherapy lacks efficacy in reducing short-term mortality in patients hospitalized with COVID-19 disease based on moderate level of evidence. We also found that the use of hydroxychloroquine in combination with azithromycin is likely associated with increased short-term mortality among COVID-19 patients.

## Data Availability

None

## Funding

This research did not receive any specific grant from funding agencies in the public, commercial, or not-for-profit sectors.

## Disclosures and conflict of interests

All authors have no financial disclosures or conflicts of interest relevant to this study.

## Notes

### Competing Interest Statement

The authors have declared no competing interest.

### Author Declarations

No IRB approval required for a systematic review of the literature

## References

1. Inglot AD. Comparison of the antiviral activity in vitro of some non-steroidal anti-inflammatory drugs. J Gen Virol 1969; 4: 203–14.

2. Chiang G, Sassaroli M, Louie M, Chen H, Stecher VJ, Sperber K. Inhibition of HIV-1 replication by hydroxychloroquine: mechanism of action and comparison with zidovudine. Clin Ther 1996; 18: 1080–92.

3. Keyaerts E, Vijgen L, Maes P, Neyts J, Van Ranst M. In vitro inhibition of severe acute respiratory syndrome coronavirus by chloroquine. Biochem Biophys Res Commun 2004; 323: 264–8.

4. de Wilde AH, Jochmans D, Posthuma CC et al. Screening of an FDA-approved compound library identifies four small-molecule inhibitors of Middle East respiratory syndrome coronavirus replication in cell culture. Antimicrob Agents Chemother 2014; 58: 4875–84.

5. Vigerust DJ, McCullers JA. Chloroquine is effective against influenza A virus in vitro but not in vivo. Influenza Other Respir Viruses 2007; 1: 189–92.

6. Farias KJS, Machado PRL, de Almeida Junior, Renato Ferreira, de Aquino AA, da Fonseca, Benedito Antônio Lopes. Chloroquine interferes with dengue-2 virus replication in U937 cells. Microbiol Immunol 2014; 58: 318–26.

7. Dowall SD, Bosworth A, Watson R et al. Chloroquine inhibited Ebola virus replication in vitro but failed to protect against infection and disease in the in vivo guinea pig model. Journal of general virology 2015; 96: 3484–92.

8. Madrid PB, Chopra S, Manger ID et al. A Systematic Screen of FDA-Approved Drugs for Inhibitors of Biological Threat Agents. PloS one 2013; 8: e60579.

9. Delvecchio R, Higa LM, Pezzuto P et al. Chloroquine, an Endocytosis Blocking Agent, Inhibits Zika Virus Infection in Different Cell Models. Viruses 2016; 8:.

10. Khan M, Santhosh SR, Tiwari M, Lakshmana Rao PV, Parida M. Assessment of in vitro prophylactic and therapeutic efficacy of chloroquine against Chikungunya virus in vero cells. J Med Virol 2010; 82: 817–24.

11. Hashem AM, Alghamdi BS, Algaissi AA, Alshehri FS, Bukhari A, Alfaleh MA, Memish ZA. Therapeutic use of chloroquine and hydroxychloroquine in COVID-19 and other viral infections: A narrative review. Travel medicine and infectious disease 2020; 35: 101735.

12. Meyerowitz EA, Vannier AGL, Friesen MGN et al. Rethinking the role of hydroxychloroquine in the treatment of COVID-19. FASEB J 2020; 34: 6027–37.

13. Miranda S, Billoir P, Damian L et al. Hydroxychloroquine reverses the prothrombotic state in a mouse model of antiphospholipid syndrome: Role of reduced inflammation and endothelial dysfunction. PLoS ONE 2019; 14: e0212614.

14. Wang M, Cao R, Zhang L et al. Remdesivir and chloroquine effectively inhibit the recently emerged novel coronavirus (2019-nCoV) in vitro. Cell Res 2020; 30: 269–71.

15. Liu J, Cao R, Xu M et al. Hydroxychloroquine, a less toxic derivative of chloroquine, is effective in inhibiting SARS-CoV-2 infection in vitro. Cell Discov 2020; 6: 16.

16. Yao X, Ye F, Zhang M et al. In Vitro Antiviral Activity and Projection of Optimized Dosing Design of Hydroxychloroquine for the Treatment of Severe Acute Respiratory Syndrome Coronavirus 2 (SARS-CoV-2). Clin Infect Dis 2020;.

17. Gao J, Tian Z, Yang X. Breakthrough: Chloroquine phosphate has shown apparent efficacy in treatment of COVID-19 associated pneumonia in clinical studies. Biosci Trends 2020; 14: 72–3.

18. Gautret P, Lagier J, Parola P et al. Hydroxychloroquine and azithromycin as a treatment of COVID-19: results of an open-label non-randomized clinical trial. Int J Antimicrob Agents 2020; 105949.

19. Vaduganathan M, van Meijgaard J, Mehra MR, Joseph J, O’Donnell CJ, Warraich HJ. Prescription Fill Patterns for Commonly Used Drugs During the COVID-19 Pandemic in the United States. JAMA: the journal of the American Medical Association 2020; 323: 2524.

20. Liberati A, Altman DG, Tetzlaff J et al. The PRISMA statement for reporting systematic reviews and meta-analyses of studies that evaluate healthcare interventions: explanation and elaboration. BMJ 2009; 339: b2700.

21. Sterne JAC, Savović J, Page MJ et al. RoB 2: a revised tool for assessing risk of bias in randomised trials. BMJ 2019; 366: 4898.

22. GA Wells, B Shea, D O’Connell, J Peterson, V Welch, M Losos, P Tugwell. The Newcastle-Ottawa Scale (NOS) for assessing the quality of nonrandomised studies in meta-analyses 2000;.

23. Guyatt GH, Oxman AD, Vist GE, Kunz R, Falck-Ytter Y, Alonso-Coello P, Schünemann HJ. GRADE: an emerging consensus on rating quality of evidence and strength of recommendations. BMJ 2008; 336: 924–6.

24. Murad MH. Clinical Practice Guidelines: A Primer on Development and Dissemination. Mayo Clin Proc 2017; 92: 423–33.

25. Chen J, Liu D, Liu L et al. [A pilot study of hydroxychloroquine in treatment of patients with moderate COVID-19]. Zhejiang Da Xue Xue Bao Yi Xue Ban 2020; 49: 215–9.

26. Peter Horby. No clinical benefit from use of hydroxychloroquine in hospitalised patients with COVID-19 — RECOVERY Trial.

27. Huang M, Tang T, Pang P et al. Treating COVID-19 with Chloroquine. J Mol Cell Biol 2020; 12: 322–5.

28. Tang W, Cao Z, Han M et al. Hydroxychloroquine in patients mainly with mild to moderate COVID-19: an open-label, randomized, controlled trial. medRxiv 2020; 2020.04.10.20060558.

29. Chen L, Zhang Z, Fu J et al. Efficacy and safety of chloroquine or hydroxychloroquine in moderate type of COVID-19: a prospective open-label randomized controlled study. medRxiv 2020; 2020.06.19.20136093.

30. Alberici F, Delbarba E, Manenti C et al. A report from the Brescia Renal COVID Task Force on the clinical characteristics and short-term outcome of hemodialysis patients with SARS-CoV-2 infection. Kidney international 2020; 98: 20–6.

31. Sánchez-Álvarez JE, Pérez Fontán M, Jiménez Martín C et al. [SARS-CoV-2 infection in patients on renal replacement therapy. Report of the COVID-19 Registry of the Spanish Society of Nephrology (SEN)]. Nefrologia 2020; 40: 272–8.

32. Geleris J, Sun Y, Platt J et al. Observational Study of Hydroxychloroquine in Hospitalized Patients with Covid-19. The New England journal of medicine 2020; 382: 2411–8.

33. A I, Da B, E H et al. Hydroxychloroquine and Tocilizumab Therapy in COVID-19 Patients -An Observational Study 2020;.

34. Kuderer NM, Choueiri TK, Shah DP et al. Clinical impact of COVID-19 on patients with cancer (CCC19): a cohort study. Lancet 2020; 395: 1907–18.

35. Magagnoli J, Narendran S, Pereira F, Cummings TH, Hardin JW, Sutton SS, Ambati J. Outcomes of Hydroxychloroquine Usage in United States Veterans Hospitalized with COVID-19. Med 2020;.

36. Mahévas M, Tran V, Roumier M et al. Clinical efficacy of hydroxychloroquine in patients with covid-19 pneumonia who require oxygen: observational comparative study using routine care data. BMJ 2020; 369:.

37. Mallat J, Hamed F, Balkis M et al. Hydroxychloroquine is associated with slower viral clearance in clinical COVID-19 patients with mild to moderate disease: A retrospective study. medRxiv 2020; 2020.04.27.20082180.

38. Paccoud O, Tubach F, Baptiste A et al. Compassionate use of hydroxychloroquine in clinical practice for patients with mild to severe Covid-19 in a French university hospital. Clin Infect Dis.

39. Rosenberg ES, Dufort EM, Udo T et al. Association of Treatment With Hydroxychloroquine or Azithromycin With In-Hospital Mortality in Patients With COVID-19 in New York State. JAMA 2020;.

40. Sbidian E, Josse J, Lemaitre G et al. Hydroxychloroquine with or without azithromycin and in-hospital mortality or discharge in patients hospitalized for COVID-19 infection: a cohort study of 4,642 in-patients in France. medRxiv 2020; 2020.06.16.20132597.

41. Singh S, Khan A, Chowdhry M, Chatterjee A. Outcomes of Hydroxychloroquine Treatment Among Hospitalized COVID-19 Patients in the United States-Real-World Evidence From a Federated Electronic Medical Record Network. medRxiv 2020; 2020.05.12.20099028.

42. Yu B, Wang DW, Li C. Hydroxychloroquine application is associated with a decreased mortality in critically ill patients with COVID-19. medRxiv 2020; 2020.04.27.20073379.

43. An MH, Kim MS, park Y et al. Treatment Response to Hydroxychloroquine and Antibiotics for mild to moderate COVID-19: a retrospective cohort study from South Korea. medRxiv 2020; 2020.07.04.20146548.

44. Chen Z, Hu J, Zhang Z et al. Efficacy of hydroxychloroquine in patients with COVID-19: results of a randomized clinical trial. medRxiv 2020; 2020.03.22.20040758.

45. Feng Z, Li J, Yao S et al. The Use of Adjuvant Therapy in Preventing Progression to Severe Pneumonia in Patients with Coronavirus Disease 2019: A Multicenter Data Analysis. medRxiv 2020; 2020.04.08.20057539.

46. Esper RB, da Silva RS, Oikawa FT et al.Empirical treatment with hydroxychloroquine and azithromycin for suspected cases of COVID-19 followed-up by telemedicine 2020;.

47. de Novales FJ, Olivencia GR, de Dios ME et al. Early Hydroxychloroquine Is Associated with an Increase of Survival in COVID-19 Patients: An Observational Study 2020;.

48. Barbosa Joshua, Kaitis Daniel, L. Kim, Lin Xinhui. Clinical outcomes of Hydroxychloroquine in Hospitalized Patients with COVID-19: A Quasi-Randomized Comparative Study 2020;.

49. Arshad S, Kilgore P, Chaudhry ZS et al. Treatment with Hydroxychloroquine, Azithromycin, and Combination in Patients Hospitalized with COVID-19. Int J Infect Dis 2020;.

50. Mikami T, Miyashita H, Yamada T, Harrington M, Steinberg D, Dunn A, Siau E. Risk Factors for Mortality in Patients with COVID-19 in New York City. J Gen Intern Med 2020;.

51. Mehra MR, Desai SS, Ruschitzka F, Patel AN. RETRACTED: Hydroxychloroquine or chloroquine with or without a macrolide for treatment of COVID-19: a multinational registry analysis. Lancet 2020;.

52. Paton NI, Lee L, Xu Y et al. Chloroquine for influenza prevention: a randomised, double-blind, placebo controlled trial. Lancet Infect Dis 2011; 11: 677–83.

53. Roques P, Thiberville S, Dupuis-Maguiraga L et al. Paradoxical Effect of Chloroquine Treatment in Enhancing Chikungunya Virus Infection. Viruses 2018; 10:.

54. De Lamballerie X, Boisson V, Reynier J et al. On chikungunya acute infection and chloroquine treatment. Vector Borne Zoonotic Dis 2008; 8: 837–9.

55. Tricou V, Minh NN, Van TP et al. A randomized controlled trial of chloroquine for the treatment of dengue in Vietnamese adults. PLoS Negl Trop Dis 2010; 4: e785.

56. Chauhan A, Tikoo A. The enigma of the clandestine association between chloroquine and HIV-1 infection. HIV Med 2015; 16: 585–90.

57. Vincent MJ, Bergeron E, Benjannet S et al. Chloroquine is a potent inhibitor of SARS coronavirus infection and spread. Virology journal 2005; 2: 69.

58. Gendelman O, Amital H, Bragazzi NL, Watad A, Chodick G. Continuous hydroxychloroquine or colchicine therapy does not prevent infection with SARS-CoV-2: Insights from a large healthcare database analysis. Autoimmun Rev 2020; 19: 102566.

59. Borba MGS, Val FFA, Sampaio VS et al. Effect of High vs Low Doses of Chloroquine Diphosphate as Adjunctive Therapy for Patients Hospitalized With Severe Acute Respiratory Syndrome Coronavirus 2 (SARS-CoV-2) Infection: A Randomized Clinical Trial. JAMA Netw Open 2020; 3: e208857.

60. K S, M L, T K et al. Hydroxychloroquine treatment of patients with human immunodeficiency virus type 1. Clin Ther 1995; 17: 622–36.

61. Perinel S, Launay M, Botelho-Nevers É et al. Towards Optimization of Hydroxychloroquine Dosing in Intensive Care Unit COVID-19 Patients. Clin Infect Dis 2020;.

62. Balevic SJ, Hornik CP, Green TP et al. Hydroxychloroquine in Patients with Rheumatic Disease Complicated by COVID-19: Clarifying Target Exposures and the Need for Clinical Trials. J Rheumatol 2020;.

63. Tleyjeh I, Kashour Z, AlDosary O et al. The Cardiac Toxicity of Chloroquine or Hydroxychloroquine in COVID-19 Patients: A Systematic Review and Meta-regression Analysis. medRxiv 2020; 2020.06.16.20132878.

64. Lane JCE, Weaver J, Kostka K et al. Safety of hydroxychloroquine, alone and in combination with azithromycin, in light of rapid wide-spread use for COVID-19: a multinational, network cohort and self-controlled case series study. medRxiv 2020; 2020.04.08.20054551.

65. Bhimraj A, Morgan RL, Shumaker AH et al. Infectious Diseases Society of America Guidelines on the Treatment and Management of Patients with COVID-19. Clin Infect Dis 2020;.

66. Wolkewitz M, Schumacher M. Survival biases lead to flawed conclusions in observational treatment studies of influenza patients. J Clin Epidemiol 2017; 84: 121–9.

67. Tleyjeh IM, Ghomrawi HMK, Steckelberg JM et al. Conclusion about the association between valve surgery and mortality in an infective endocarditis cohort changed after adjusting for survivor bias. J Clin Epidemiol 2010; 63: 130–5.

68. Fewell Z, Davey Smith G, Sterne JAC. The Impact of Residual and Unmeasured Confounding in Epidemiologic Studies: A Simulation Study. Am J Epidemiol 2007; 166: 646–55.

